# Non-Invasive Scale Measurement of Cardiac Output Compared with the Gold-Standard Direct Fick Method: A Feasibility Study

**DOI:** 10.1101/2021.03.28.21254243

**Authors:** Daniel Yazdi, Sarin Patel, Suriya Sridaran, Evan Wilson, Sarah Smith, Corey Centen, Leah Gillon, Sunil Kapur, Julie A. Tracy, Katherine Lewine, David M. Systrom, Calum A. MacRae

## Abstract

**Background:** Objective markers of cardiac function are limited in the outpatient setting and may be beneficial for monitoring patients with chronic cardiac conditions.

**Objective:** We assess the accuracy of a scale, with the ability to capture ballistocardiography, electrocardiography, and impedance plethysmography signals from a patient’s feet while standing on the scale, in measuring stroke volume and cardiac output compared to the gold-standard direct Fick method.

**Methods:** Thirty-two patients with unexplained dyspnea undergoing level 3 invasive cardiopulmonary exercise test at a tertiary medical center were included in the final analysis. We obtained scale and direct Fick measurements of stroke volume and cardiac output before and immediately after invasive cardiopulmonary exercise test.

**Results:** Stroke volume and cardiac output from a cardiac scale and the direct Fick method correlated with r = 0.81 and r = 0.85, respectively (P < 0.001 each). The mean absolute error of the scale estimated stroke volume was -1.58 mL, with a 95% limits of agreement (LOA) of -21.97 mL to 18.81 mL. The mean error for the scale estimated cardiac output was -0.31 L/min, with a 95% LOA of -2.62 L/min to 2.00 L/min. The change in stroke volume and cardiac output before and after exercise were 78.9% and 96.7% concordant, respectively between the two measuring methods.

**Conclusions:** This novel scale with cardiac monitoring abilities may allow for non-invasive, longitudinal measures of cardiac function. Using the widely accepted form factor of a bathroom scale, this method of monitoring can be easily integrated into a patient’s lifestyle.

## Introduction

Cardiovascular disease, in particular heart failure (HF), is a major health and economic problem worldwide, expected to increase in incidence and prevalence due to the aging population and rise in co-morbidities ^1,2^. Novel approaches for easily monitoring cardiac function trends over time in the home environment may prove to be important in dealing with these conditions. Accelerated by the COVID-19 pandemic, the field of medicine is increasingly shifting towards telemedicine and remote patient monitoring, welcoming innovation ^3,4^. In this study, we investigate the accuracy of a connected cardiac scale with ballistocardiography (BCG), impedance plethysmography (IPG), and electrocardiography (ECG) sensors in measuring stroke volume and cardiac output compared to the direct Fick method.

BCG measures the effects of the cyclical hemodynamic forces transmitted from the heart with each cardiac systolic ejection ^5^. The methodology for BCG was developed and popularized in the 1950s, but its use waned later in the century due to the impractical nature of the apparatus, limited reliability of the measurements in diseased states, and a focus on other measures of cardiovascular function, such as blood pressure recordings ^6,7^. However, over the past decade BCG has regained popularity as a result of the ability to obtain measurements from novel sensor platforms, such as bathroom scales, advances in data processing and machine learning algorithms, and the emergence of more rigorous studies demonstrating the utility of BCG recordings from patients with cardiac diseases ^8–10^. IPG measures pulsatile blood flow via changes in electrical impedance. This measurement, along with the ECG, can identify important cardiac time intervals such as valvular opening and closing, measures of contractility such as left ventricular ejection time, and estimates for stroke volume and cardiac output ^11^. The simultaneous extraction of ECG, IPG, and BCG signals from a single scale measurement can enhance the amount of cardiovascular information obtained for any individual signal in isolation.

In this study, Bodyport Inc., a company based in San Francisco, CA, and the Brigham and Women’s Hospital collaborated to assess the accuracy of the Bodyport Cardiac Scale in non-invasively measuring stroke volume and cardiac output. The cardiac scale has the form of a bathroom scale and has a multi-sensor system that can capture a single lead ECG, IPG, and BCG signals when a patient stands on the scale (Figure 1). This is the first study to our knowledge to investigate the accuracy of such a scale in measuring stroke volume and cardiac output compared to the gold-standard direct Fick method from pulmonary arterial catheters. Our findings demonstrate a robust correlation and relatively accurate mean error between direct Fick measurements of stroke volume and cardiac output and those obtained from the cardiac scale. These data support the potential clinical utility of such home-based sensors.

**Figure 1:**
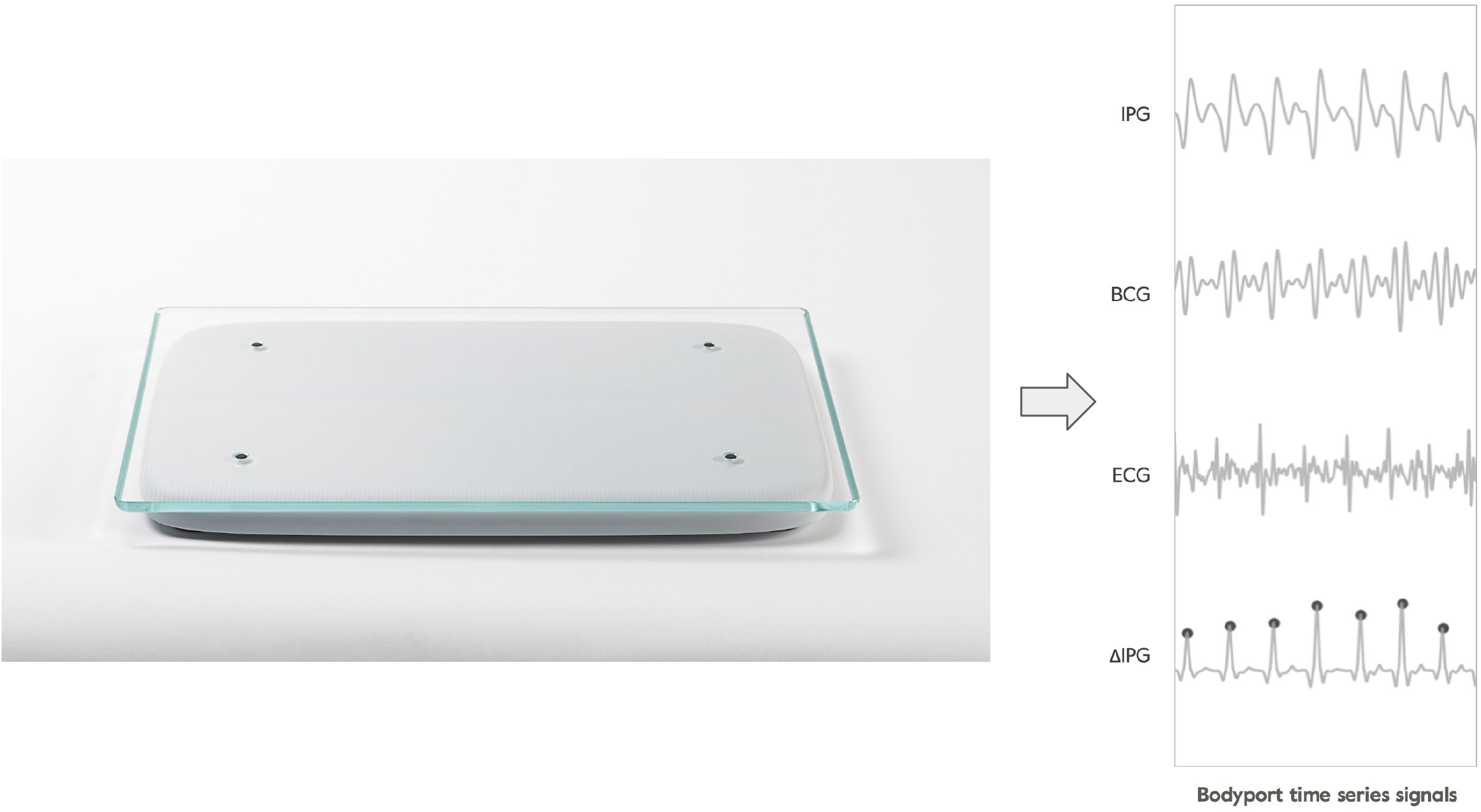
Left: Image of the Bodyport Cardiac Scale. Right: Example of scale-derived signals for the impedance plethysmograph (IPG), ballistocardiograph (BCG), electrocardiograph (ECG), and derivative of the IPG (ΔIPG).

## Methods

### Subject Population

Fifty-six subjects undergoing a level 3 invasive cardiopulmonary exercise test (iCPET) at the Shapiro Cardiovascular Center at Brigham and Women’s Hospital were recruited and consented between July 2018 and January 2019. The patients were each undergoing the iCPET to evaluate unexplained dyspnea. These patients had an array of underlying comorbidities including heart failure, pulmonary artery hypertension, peripheral vasomotor abnormalities, valvular pathologies, and pulmonary diseases. The study was not designed to discriminate the accuracy of the cardiac output measurements dependent on the patient’s underlying medical condition. Of the 56 subjects enrolled in the study, 32 subjects (9 male, 23 female) were included in the final analysis. Twenty-four subjects were excluded from the final analysis. Among the 24 patients excluded, eleven subjects experienced light-headedness at the completion of the iCPET making it unsafe for them to stand on the scale. Six subjects had missing or incorrect reference data due to instrumentation related issues (i.e. patient pulled face mask off before Fick measurement completed). Seven patients had poor balance during the post-exercise recovery measurement requiring assistance from the clinical staff, which caused excessive motion artifacts requiring exclusion of the data. The age range for participants was 26-78 years (mean 51.7 years, SD 14.5 years).

### Study Protocol

We obtained scale measurements before and immediately after the iCPET. Ultrasound guided pulmonary artery catheters were placed prior to patient arrival in the iCPET lab. Once in the iCPET lab, patients were asked to stand, and a baseline pulmonary artery measurement was obtained followed immediately by a 2-minute baseline measurement on the cardiac scale.

Patients then mounted an upright cycle ergometer to perform the exercise portion of the test. The exercise workload was gradually increased in a ramped protocol until the patient reached exhaustion or developed objective evidence of hemodynamic instability or myocardial ischemia. After the exercise limit was reached, patients dismounted from the bicycle as quickly as possible to obtain a recovery pulmonary artery catheter measurement. This was immediately followed by a 2-minute recovery cardiac scale measurement.

### Direct Fick Measurements

The measurements obtained from the metabolic cart, radial and pulmonary artery catheters before and immediately after the iCPET study include ventilation, pulmonary gas exchange, venous and arterial blood gases, a 12-lead ECG, heart rate, pulmonary artery pressure, blood pressure from a radial artery catheter. Cardiac output was calculated using the direct Fick method as the current gold standard. Stroke volume was simply derived from its relationship with the measured cardiac output and heart rate.

### Cardiac Scale Measurements

The Bodyport Cardiac Scale is a physical platform on which the patient stands with bare feet. Using embedded sensors, the scale measures three cardiovascular biosignals that are used to extract various cardiac biomarkers. The BCG, ECG, and IPG signals are analyzed by Bodyport’s software and proprietary algorithms to extract characteristic features in each waveform that are used to estimate parameters including heart rate, heart rate variability, cardiac time intervals, and signal morphological features used to derive estimates of stroke volume and cardiac output.

### Feature Selection

We developed a feature set derived from the BCG, ECG, and IPG signals from the combined pre-exercise and post-exercise measurements to function as inputs in the model development for the estimation of stroke volume and cardiac output. The feature set included time intervals and amplitudes extracted from time aligned ensemble averaged signals. Certain features were chosen based on existing equations used in impedance cardiography and ballistocardiography, including those of Kubicek and Starr ^12^. The BCG J-wave amplitude, which correlates with pulse pressure, along with additional proprietary electromechanical parameters from the BCG, ECG, and IPG signals, specifically pre-ejection period and left ventricular ejection time, were used to develop the final feature set ^5^. Heart rate was used as a correction factor applied to time interval features. Additional non-cardiovascular parameters collected directly from the scale, such as body weight and basal impedance, were also incorporated into the model to remove the need for calibration and compensate for individual anthropomorphic variability.

### Data Processing and Analysis

The scale signals (BCG, ECG, IPG) were filtered and interpolated prior to feature extraction and model validation. For this calibration exercise, signal regions containing motion artifacts or excessive noise were objectively identified and removed. Motion artifacts and other sources of signal interference were detected through adaptive thresholding and monitoring of the patient’s center of pressure during the measurement. Linear phase digital lowpass and highpass filters were applied to the signals to prevent distortion. Cutoff frequencies were established independently for each signal and ranged from 0.5Hz to 50Hz. All three signals were simultaneously sampled at 250Hz. Ensemble averaged waveforms were constructed from the real-time signals. Specific features were identified on the averaged waveforms using proprietary algorithms optimized for the Bodyport device. These features included signal amplitudes, such as the BCG J-wave magnitude, and temporal relationships between each of the averaged waveforms, such as pre-ejection period and other systolic time intervals.

Regression model training for stroke volume was accomplished using a Tree-based Pipeline Optimization Tool (TPOT) ^13^. This tool evaluates the performance of individual regression models, while optimizing hyperparameters. The following models were preselected to be used by TPOT: Linear Regression, Lasso, Elastic Net, Ridge, Random Forest, Support Vector Regression, Multi-Layer Perceptron. Feature preprocessors were also predefined to be included as part of the TPOT process. TPOT iterates over combinations of models and preprocessors, as well as the hyperparameter space. Each trained model in this stage used K-Fold cross-validation using three folds. Mean absolute error was used to optimize model training accuracy. The final model used an ensemble regression pipeline consisting of a random forest and gradient boosting regression and was then evaluated using leave-one-out cross-validation. This cross-validation technique fits the model on all but one measurement, which is then used as the test measurement. Each measurement was held out once and the final accuracy was determined based on the performance of all held out test measurements. Bodyport-derived heart rate and stroke volume were used to derive an estimate of cardiac output. The p-value for the correlation coefficients was calculated using the Wald test with t-distribution of the test statistic. The Bland-Altman limits of agreement analysis for the combined pre-exercise and post-exercise data set accounted for the multiple measurements from the same subject using the methodology described by Bland and Altman^14^.

## Results

The multivariate regression model demonstrated a strong relationship between the scale and Fick-derived estimates for stroke volume and cardiac output (Figure 2). The stroke volume in the pre-exercise analysis correlated with the Fick derived stroke volume with a coefficient of r = 0.77 (P < 0.001), mean error (bias) of -2.08 mL, standard deviation (SD) of 10.65 mL, 95% limits of agreement (LOA) for the mean error of -22.95 mL to 18.79 mL, and percentage error (PE), defined as the 95% LOA (± 1.96 SD) divided by the average Fick derived stroke volume, of 39.14% (Table 1). In the post-exercise analysis, the model had a correlation coefficient of r = 0.84 (P < 0.001), mean error of -1.09 mL, SD of 9.92 mL, 95% LOA of -20.53 mL to 18.35 mL, and a PE of 33.66%. The combined pre-exercise and post-exercise data set had a correlation coefficient of r = 0.81 (P < 0.001), mean error -1.58 mL, 95%, SD of 10.30 mL, LOA of -21.97 mL to 18.81 mL, and PE of 36.73%.

**Table 1.** Summary statistics and accuracy metrics for scale estimates of stroke volume and cardiac output compared to the direct Fick method.

|  | Scale measure<br>Mean (Range) | Fick measure<br>Mean (Range) | Correlation<br>coefficient (r) | Mean error,<br>95% LOA | Percentage<br>error |
| --- | --- | --- | --- | --- | --- |
| **Stroke Volume** |  |  |  |  |  |
| Pre-exercise | 51.23 mL<br>(33.72-75.77) | 53.31 mL<br>(29.0-89.3) | 0.77 | -2.08 mL,<br>(-22.95, 18.79) | 39.14% |
| Post-exercise | 56.65 mL<br>(30.25-88.62) | 57.74 mL<br>(24.8-95.4) | 0.84 | -1.09 mL,<br>(-20.53, 18.35) | 33.66% |
| Combined | 53.94 mL<br>(30.25-88.63) | 55.52 mL<br>(24.8-95.4) | 0.81 | -1.58 mL,<br>(-21.97, 18.81) | 36.73% |
| **Cardiac Output** |  |  |  |  |  |
| Pre-exercise | 4.39 L/min<br>(3.07-6.74) | 4.56 L/min<br>(2.77-8.13) | 0.70 | -0.17 L/min,<br>(-1.97, 1.63) | 39.52% |
| Post-exercise | 6.20 L/min<br>(4.20-9.86) | 6.65 L/min<br>(3.45-13.07) | 0.83 | -0.45 L/min,<br>(-3.11, 2.20) | 39.89% |
| Combined | 5.29 L/min<br>(3.07-9.85) | 5.61 L/min<br>(2.77-13.07) | 0.85 | -0.31 L/min,<br>(-2.62, 2.00) | 41.18% |

**Figure 2:**
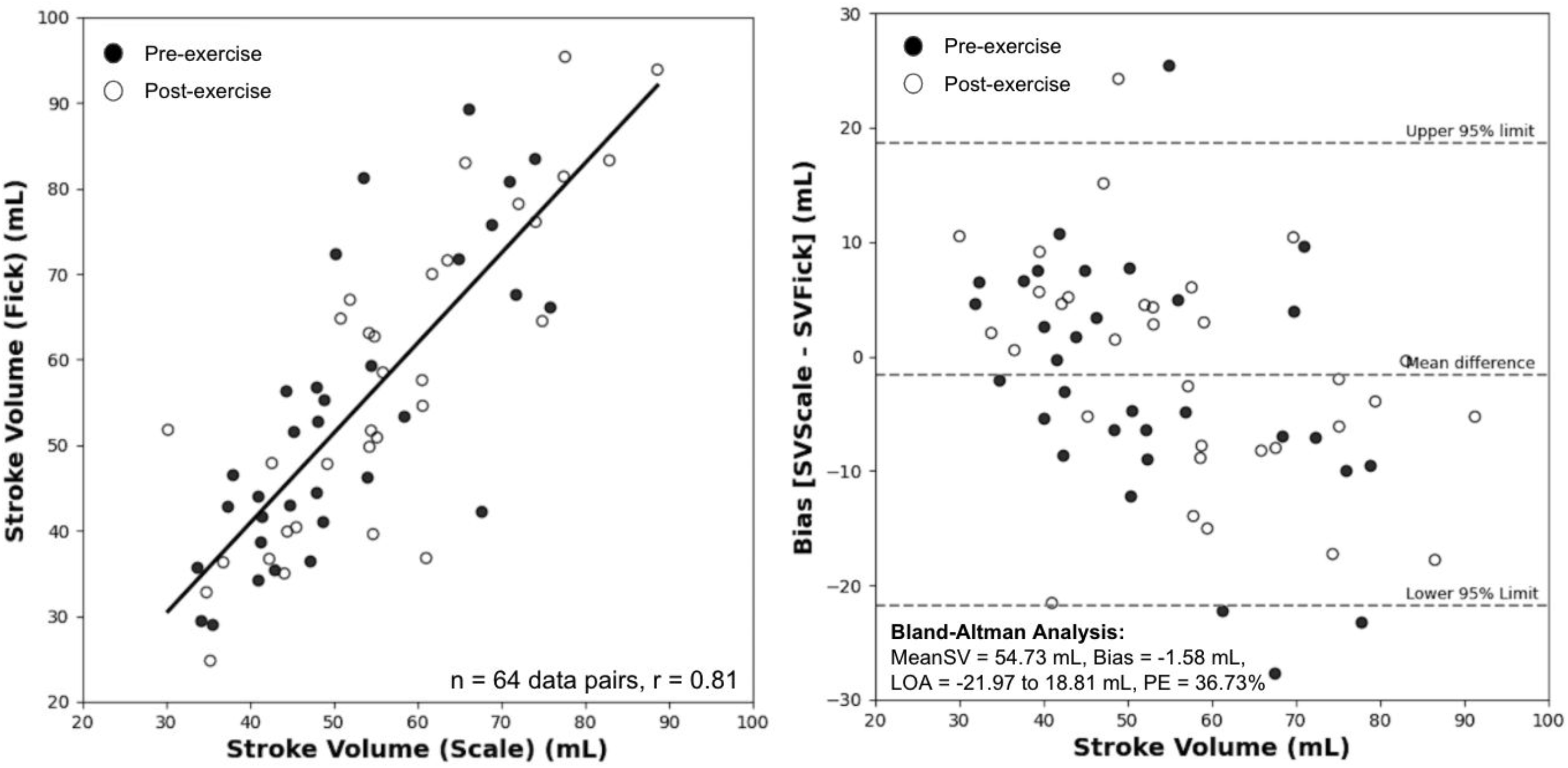
Left: Scatter plot with regression line for stroke volume measured by the scale and direct Fick method (64 data pairs, r = 0.81). Pre-exercise data denoted with a black circle and post-exercise data with a white circle. Right: Bland-Altman analysis with mean error (bias) of -1.58 mL, 95% LOA of -21.97 to 18.81 mL, PE = 36.73%. LOA = limits of agreement, MeanSV = Mean stroke volume for combined scale and Fick measurements, PE = percentage error.

Cardiac output was estimated using the scale-derived stroke volume and heart rate. In the pre-exercise set, the correlation coefficient was r = 0.70 (P < 0.001) (Figure 3), mean error of -0.17 L/min, SD of 0.92 L/min, 95% LOA -1.97 L/min to 1.63 L/min, and a PE of 39.52%. The post-exercise set had correlation coefficient was r = 0.83 (P < 0.001), mean error of -0.45 L/min, SD of 1.36 L/min, 95% LOA of -3.11 L/min to 2.2 L/min, and PE of 39.89%. Combining the two sets yielded a correlation coefficient of r = 0.85 (P < 0.001), mean error of -0.31 L/min, SD of 0.98 L/min, 95% LOA -2.62 L/min to 2.00 L/min, and PE of 41.18%.

**Figure 3:**
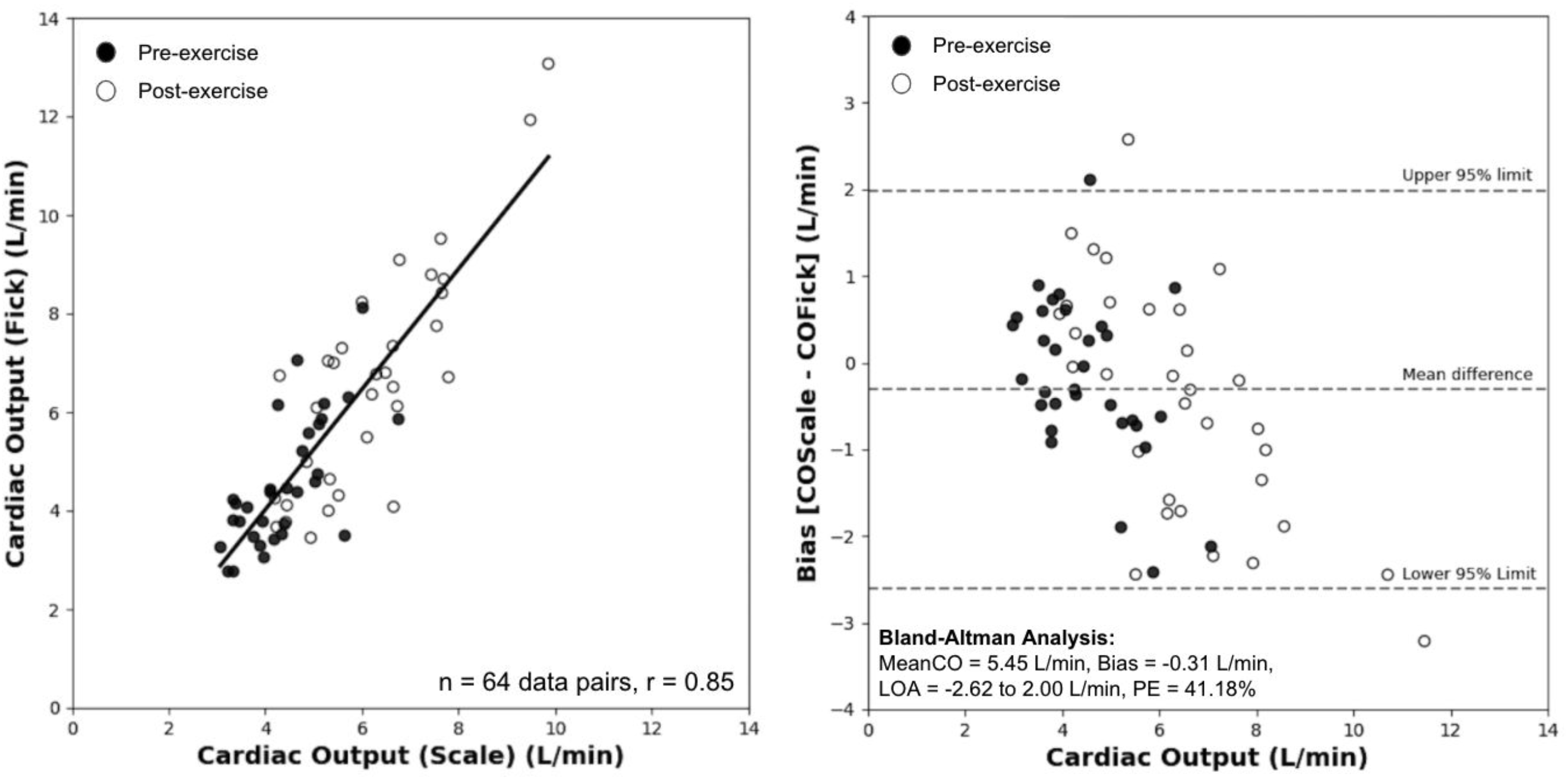
Left: Scatter plot with regression line for cardiac output measured by the scale and direct Fick method (64 data pairs, r = 0.85). Pre-exercise data denoted with a black circle and post-exercise data with a white circle. Right: Bland-Altman analysis with mean error (bias) of -0.31 mL, 95% LOA of -2.62 to 2.00 L/min, PE = 41.18%. LOA = limits of agreement, MeanCO = Mean cardiac output for combined scale and Fick measurements, PE = percentage error.

The change in stroke volume and cardiac output were evaluated pre- and post-exercise test for both the scale-derived and Fick methods, to assess if the scale can trend directional changes in these perfusion markers. We measured the degree of agreement between these two methods by calculating their concordance: the fraction of patients for which the change in stroke volume or cardiac output (post-exercise minus pre-exercise) were either both positive or negative using both modalities. The concordance for stroke volume was 78.9% and 96.7% for cardiac output (P < 0.001, each, derived from the concordance correlation) (Figure 4). To reduce statistical noise from the analysis, we excluded data points (13 for stroke volume and 2 for cardiac output) where the change in stroke volume or cardiac output was less than 15% of the mean value in the study ^15^.

**Figure 4:**
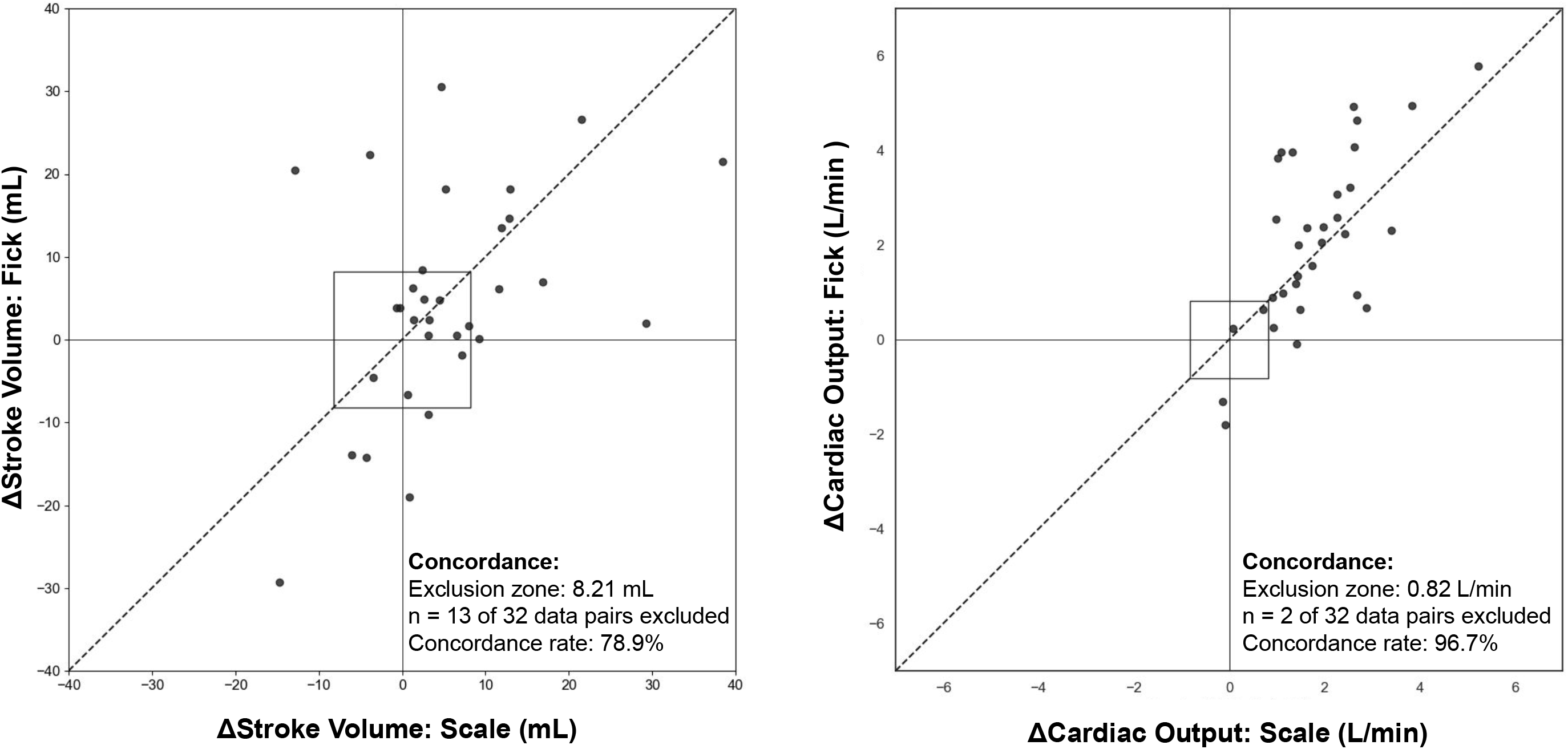
Concordance plot: Left: Change in stroke volume post-exercise minus pre-exercise from the scale vs the direct Fick method. Right: Similar plot for cardiac output. A central exclusion zone (square) represents the data within 15% of the mean stroke volume or cardiac output in the study, as they contain a high level of random variation compared to changes in the cardiac output. The line of identity y = x is shown.

After exercise, the average cardiac output increased by 1.81 L/min when measured by the scale (4.39 L/min pre-exercise to 6.20 L/min post-exercise), and by 2.09 L/min (from 4.56 L/min to 6.65 L/min) when measured by the Fick method. The heart rate increased by an average of 24 beats per minute (bpm) (88 bpm pre-exercise to 112 bpm post-exercise) on the scale, and 40 bpm (76 bpm pre-exercise to 116 bpm post-exercise) for the Fick measurement. The stroke volume increased by an average of 5.24 mL (51.23 mL pre-exercise to 56.65 mL post-exercise) on the scale and 4.43 mL (53.31 pre-exercise mL to 57.74 mL post-exercise) using the Fick method.

## Discussion

### Principal Findings

The results demonstrate a strong relationship between the cardiac scale and direct Fick estimates for stroke volume and cardiac output. The correlation persisted through a dynamic range of stroke volumes (30-90 mL) and cardiac outputs pre- and post-exercise (3-10 L/min). The mean error for the cardiac output of -0.31 L/min is less than other non-invasive measures of cardiac output, such as doppler ultrasound and bioimpedance, which have errors of approximately 0.8 L/min and 0.6L/min, respectively, compared to thermodilution and Fick in a meta-analysis^16^. In another meta-analysis with 26 patients, thermodilution versus direct Fick had a mean error of 2.3 L/min, highlighting the significant variability in cardiac output measurement techniques ^17^. One of the primary advantages of the scale is to provide granular longitudinal measures of cardiac perfusion in the home setting, not to entirely replace the “gold-standard” catheterization laboratory measurement. Furthermore, stroke volume and cardiac output can be combined with other metrics obtained from the BCG, ECG, and IPG sensors to create a more robust model for overall cardiovascular hemodynamic status.

The stroke volume and cardiac output concordance before and immediately after exercise were 78.9% and 96.7%, respectively. Current advice for trending ability in cardiac output studies is a concordance greater than 92%, which is achieved in this cardiac output analysis but not the stroke volume ^18^. The increased concordance for the cardiac output is likely a result of an increased heart rate post-exercise, compared to the more variable response seen in the stroke volume. Because exercise is incorporated in the iCPET, cardiac output is expected to increase. This is why a majority of the data points fall in the upper right corner of the cardiac output concordance plot (Figure 4). Another experimental design is necessary to better detect reductions in cardiac output.

Having all sensors integrated in one device is imperative for the future adoption of such a technology, since applying multiple sensor technologies simultaneously is cumbersome and would likely yield low patient adherence. The analysis of these orthogonal sensor signals is further enhanced by ongoing advancements in signal processing and machine learning techniques. Utilizing the form factor of a bathroom scale will also enhance patient adherence, since taking scale measurements is a behavior already adapted by many patients, especially those with cardiac conditions such as heart failure.

A non-invasive, scalable, and inexpensive method for assessing cardiac function could have widespread applications in medicine. Robust estimates of stroke volume and cardiac output may help monitor the cardiac performance of patients with chronic conditions, such as heart failure, and facilitate early detection of decompensation and the virtual titration of goal-directed medical therapy. Given the simplicity of use and existing user behaviors for self-weighing on a scale, longitudinal data can be obtained and trended in large populations to identify novel biomarkers of cardiovascular health.

### Limitations

This proof-of-concept study was not designed or powered to measure changes in cardiac output in relation to the patients’ underlying medical conditions. The 95% LOA in cardiac output during the post-exercise phase was up to 3 L/minute. This is partially accounted for by the 2-minute time delay between the Fick and scale measurements, allowing interval cardiac recovery. There was a decrease in post-exercise heart rate by an average of 16 bpm between the Fick and scale measurements, while the stroke volume only decreased by an average of 1.09 mL. With an average stroke volume of 57.74 mL in the post-exercise Fick subgroup, a decrease in heart rate by 16 bpm would reduce the cardiac output by 0.92 L, supporting a post-exercise cardiac output 95% LOA closer to 2 L/min, similar to that seen in the pre-exercise analysis. Additional sources of error include patient balance post exertion, the study’s limited sample size, and intrinsic errors of both the direct Fick measurement and scale technology. The exclusion of patients in this study was largely due to unstable hemodynamics (hypotension), symptomatic factors during testing (dizziness, poor balance), and incomplete reference measurements. The application of this scale in its normal use, a stable, non-stress setting such as the patient’s home, would mitigate these factors.

## Conclusions

This study highlights how enhancements in technology have allowed for the integration of BCG, ECG, and IPG sensors into form factors (e,g, a bathroom scale creating novel methods for assessing cardiac function. The combination of the BCG, ECG, and IPG signals provides important electromechanical cardiac information that enhances the estimations of stroke volume and cardiac output compared to the IPG or BCG signal alone. We observed strong correlations between scale-derived and Fick-derived estimates of stroke volume and cardiac output, with r = 0.81 and 0.85, respectively. The mean errors of the scale estimates for stroke volume and cardiac output were relatively accurate at -1.58 mL (−21.97, 18.81 mL) and -0.31 L/min (−2.62, 2.00 L/min), respectively. The scale and direct Fick estimates for cardiac output were strongly concordant pre- and post-exercise at 96.7%, demonstrating the ability for scale to trend increases in cardiac output. Future studies will gather additional data to improve the model and will also assess longitudinal scale measurements from individuals with in clinical settings to better understand how these biomarkers, when integrated, can be used to assess relevant changes in a range of disorders.

## Data Availability

The data sets generated during and/or analyzed during the current study are not publicly available due their proprietary nature but are available from the corresponding author on reasonable request.

## Acknowledgements

We would like to thank the staff of the Brigham and Women’s Hospital cardiopulmonary exercise lab for helping collect the data.

## Conflicts of Interest

Daniel Yazdi and Sarin Patel are employees at Bodyport Inc. Sarah Smith and Corey Centen are founders of Bodyport Inc. Calum A. MacRae is an advisor for Bodyport Inc.

## Abbreviations

BCG: ballistocardiography
ECG: electrocardiography
HF: heart failure
iCPET: invasive cardiopulmonary exercise test
IPG: impedance plethysmography
LOA: limits of agreement
PE: percentage error
TPOT: Tree-based Pipeline Optimization Tool

## Notes

### Funding Statement

No external funding was received for this study.

### Author Declarations

Approved by Brigham and Women's Hospital IRB.

